# Reported autism diagnosis strongly predicts psychotic-like experiences in the Adolescent Brain Cognitive Development cohort

**DOI:** 10.1101/2020.02.07.20021170

**Authors:** Amandeep Jutla, Meghan Rose Donohue, Jeremy Veenstra-VanderWeele, Jennifer H. Foss-Feig

## Abstract

Although the schizophrenia rate is increased in autism spectrum disorder (ASD), it is difficult to identify which youth with ASD will go on to develop psychosis. We explored the relationship between ASD and emerging psychotic-like experiences in the Adolescent Brain Cognitive Development (ABCD) cohort of school-aged children. We predicted that ASD would robustly predict psychotic-like experience severity, even relative to other established predictors, and that ASD youth with psychotic-like experiences would have a characteristic neurocognitive profile. In a sample of 9,130 youth aged 9–11, we fit regression models that included parent-reported ASD, family history of psychosis, lifetime trauma, executive function, processing speed, working memory, age, sex, race, ethnicity, and income-to-needs ratio as predictors of Prodromal Questionnaire – Brief Child (PQ-BC) score. We assessed cognitive profiles using analysis of variance on NIH Toolbox measures. ASD increased PQ-BC distress scores by 2.46 points (95% CI 1.32 – 3.60), an effect at least as large as those of family history of psychosis (1.05 points, 95% CI 0.56 – 1.53), Latinx ethnicity (0.99 points, 95% CI 0.52 – 1.45) and black race (0.89 points, 95% CI 0.30 – 1.48). We did not identify a unique cognitive profile for ASD youth with psychotic-like experiences. Our finding that ASD predicts psychotic-like symptoms in youth is consistent with previous literature, and adds new information in suggesting that ASD is a strong predictor of psychotic-like experiences even when compared to other established SCZ risk factors.

## 1 Introduction

Autism spectrum disorder (ASD) and schizophrenia (SCZ) are separate diagnostic entities that in some respects show clear divergence. ASD, typically diagnosed in childhood, is characterized by the combined presence of social communication deficits and restricted or repetitive behaviors^1^ and tends either to have a stable course or to show clinical improvement with time^2^. SCZ, typically diagnosed in late adolescence or early adulthood^3^, is characterized by psychosis, which reflects a loss of contact with reality, and manifests as delusional beliefs or hallucinatory experiences^4^. Unlike ASD, it tends to be associated with progressive functional and cognitive decline^5^.

Despite their clear differences, ASD and SCZ also have notable similarities. Both have an impact on social skills, with impaired social communication a core deficit in ASD and an important cause of long-term disability in SCZ^6^. Both are characterized by deficits in non-social cognition, sharing impairments in executive function^7,8^, processing speed^9,10^ and working memory^11,12^.

Consistent with this, genome-wide association data demonstrate robust shared risk between ASD and SCZ^13^. Recent meta-analytic findings suggest that ASD diagnosis is in fact a SCZ risk factor. Individuals with ASD appear to be 3 to 4 times more likely to develop SCZ than members of the general population^14,15^.

Whether ASD is a more general risk factor for childhood psychotic-like experiences, in addition to a specific risk factor for SCZ diagnosis, remains an open question. “Psychotic-like experiences” constitute psychotic symptoms such as delusions or hallucinations in the absence of a frank psychotic disorder^16^, and in children may have prognostic value. For example, early psychotic-like experiences at age 11 increase the risk of schizophreniform disorder, a precursor to SCZ, at age 26^17^. In recent years, as SCZ has increasingly been viewed through a neurodevelopmental lens^3^, psychotic-like experiences have become a focus of research in their own right, as they may present an opportunity for intervention before clinical SCZ is diagnosed^18^.

When psychotic-like experiences persist and are associated with distress or impairment, they are thought to represent a meaningful diagnostic entity known as the “clinical high-risk for psychosis (CHR) syndrome,” sometimes referred to as “attenuated” or “prodromal” psychosis^19^. Even though the CHR syndrome progresses to frank SCZ in just one-fourth of cases within 36 months of its identification^20^, CHR youth who do not progress continue to have pronounced social and cognitive impairment^21^ and may benefit from specialized attention.

Recent findings suggest that ASD youth with CHR show a symptom pattern and rate of conversion to psychosis similar to the general CHR population, yet may be under-referred to CHR clinical settings^22^. This may in part stem from the dearth of research exploring the nature and extent of the relationship between ASD and early psychotic-like experiences.

In particular, the effect size of ASD relative to other factors thought to predict SCZ or psychotic-like experiences, such as family history of psychosis^23,24^, childhood trauma^25^, or race^26,27^, is unknown. It is also unclear whether ASD children with psychotic-like experiences show a pattern of impairment in executive function, working memory and processing speed dissociable from the patterns seen in those with ASD alone or psychotic-like experiences alone in this age group.

We sought to address these gaps in knowledge by investigating ASD and psychotic-like experiences in a large national sample of 9-to 11-year-olds. We hypothesized that (1) an ASD diagnosis would predict severity of psychotic-like experiences, and (2) the profile of executive function, working memory, and processing speed impairment would differ among ASD children with psychotic-like experiences, ASD children without psychotic-like experiences, and children with psychotic-like experiences but not ASD.

## 2 Method

### 2.1 Study sample

Our study drew from the Adolescent Brain Cognitive Development (ABCD; RRID:SCR_015769) study cohort. Detailed descriptions of the ABCD study’s design and recruitment strategy have been published elsewhere^28,29^, but in brief, the study recruited 11,875 children from 21 sites across the United States. Recruitment used probability sampling of schools, with a goal of obtaining a representative cross-section of U.S. youth aged 9 to 11 to follow longitudinally over ten years into early adulthood. The ABCD study was approved by the institutional review board at each participating site.

Our study used baseline cross-sectional data collected from each participant between September 2016 and October 2018. Given our aims, we excluded 2,745 ABCD participants who were missing data for any of the following variables: ASD diagnosis, psychotic-like experience rating, family history of psychosis, age-adjusted standard scores for NIH toolbox measures of cognition, trauma history, age, sex, race, ethnicity, household income category, number of people in the household, history of speech delay, maternal age at birth, and paternal age at birth (see **Table S1** for missingness by variable). This yielded a sample of 9,130 youth.

### 2.2 Assessment measures

#### ASD

As the ABCD study did not collect ASD-specific measures at baseline, we ascertained ASD based on whether a participant’s parent(s) reported an existing ASD diagnosis during the screening interview. Note that we interrogated the validity of this ascertainment method by estimating a logistic regression model, ancillary to our main analysis, in which we examined whether variables known to be predictive of an ASD diagnosis (including speech delay, male sex, and older parental age) predicted parent report of ASD diagnosis during screening. Model coefficients are reported in the results section below.

#### Psychotic-like experiences

We identified psychotic-like experiences using the Prodromal Questionnaire - Brief Child version (PQ-BC). The PQ-BC is an adapted version of the Prodromal Questionnaire - Brief (PQ-B), a 21-item screening questionnaire for psychotic-like experiences in adolescents and adults. Each PQ-B item asks about the presence of a psychotic-like experience and then has the respondent rate, on a five-point scale, how much distress the symptom causes if present. The PQ-BC retains the PQ-B’s 21 items, but with simplified, child-appropriate wording. To ensure that children understand the questions posed, these items are administered as an interview rather than a questionnaire, with the distress scale paired with a visual response analog^18,30^.

The PQ-BC, like the PQ-B, is scored by deriving two indices. For the “total” score, ranging from 0 to 21, one point is assigned per symptom endorsed. For the “distress” score, ranging from 0 to 105, 1 to 5 points are assigned per symptom endorsed, based on a distress rating (where 1 indicates “no distress” and 5 “severe distress”)^18,30^. As psychotic-like experiences are thought to be more predictive of CHR when associated with distress,^31^ our study focused on the PQ-BC distress score.

We analyzed the distress score in two ways: 1) by using the score itself as a continuous indicator of psychotic-like experience severity, and 2) by using an empirical cutoff of a score 2 standard deviations above the mean to represent the binary presence of “likely significant” psychotic-like experiences. We used this cutoff because no studies have yet recommended a meaningful PQ-BC cutoff, and the applicability of recommendations from previous studies using the PQ-B as a screening tool^30,31^ to a younger population is not clear.

#### Family history of psychosis

We defined family history of psychosis as a response of “yes” to the question “Has any blood relative of your child ever had a period lasting six months when they saw visions or heard voices or thought people were spying on them or plotting against them?”

#### Cognitive measures

The ABCD cognitive assessment^32^ included three NIH Toolbox Cognition Battery instruments relevant to our study: Dimensional Change Card Sort (RRID:SCR_003616),^33,34^, Pattern Comparison Processing Speed (RRID:SCR_003623)^35^, and List Sorting Working Memory (RRID:SCR_003626) [tulskyNIHToolboxCognition2013]. These are continuous measures of executive function, processing speed, and working memory respectively. All three have shown adequate convergent and discriminant validity, as well as good reliability in children, with reported test-retest intraclass correlation coefficients of 0.86 – 0.95 for Dimensional Change Card Sort^34^, 0.75 – 0.90 for Pattern Comparison Processing Speed^35^, and 0.78 – 0.91 for List Sorting Working Memory^36^.

#### Trauma history

Each participant’s parent(s) received the traumatic events section of the Kiddie Schedule for Affective Disorders and Schizophrenia Present and Lifetime version (K-SADS-PL) structured interview^37^. This entails a series of yes/no questions about whether the participant has ever had certain lifetime traumatic experiences. Using these data, we derived a continuous “trauma index” for each participant, representing the number of traumatic experiences endorsed.

#### Demographic variables

Of the key demographic variables available in the ABCD dataset^38^, we included age at evaluation and sex assigned at birth as reported. Small numbers of participants for racial groups other than white or black required us to create an “other” racial group for statistical analysis. This group included Chinese, Filipinx, Indian, Japanese, Korean, Vietnamese, Native American, Native Alaskan, Native Hawaiian, Guamanian, Samoan, “other Asian,” and “other Pacific Islander” participants, as well as “other race,” “unknown race,” and multiracial participants. We included ethnicity as a binary variable (Latinx vs. non-Latinx) independent from race.

We derived an income-to-needs ratio for each participant by (1) transforming their reported household income category into the mean income for that category and (2) dividing this by the poverty threshold for that participant’s family size in the year that participant was interviewed^39^.

#### Other variables

We used a reported history of speech delay (defined as a parent response of “later” to the question “Would you say his/her speech development was earlier, average, or later than most other children?”), maternal age at birth, and paternal age at birth in our ancillary analysis of ASD predictors.

### 2.3 Statistical approach

First, we compared participants with and without ASD across the variables described above. For continuous variables, we compared means with Welch’s two-sided unequal variances *t*-test, and for categorical variables we compared frequency counts with Pearson’s χ^2^. Within each of these sets, we used the Benjamini-Hochberg procedure to control the false discovery rate.

Next, to ascertain whether ASD diagnosis predicted psychotic-like experiences (hypothesis 1), we estimated two regression models. The first model was a linear regression against continuous PQ-BC distress score. The second was a binary logistic regression against a PQ-BC distress cutoff ≥ 2 standard deviations above the mean. Both models had a multilevel (“mixed-effect”) structure, with family unit and ABCD recruitment site as nested random effects^40^, and were fit by restricted maximum likelihood using bound optimization by quadratic approximation.

Both models included the following inputs as fixed effects: ASD diagnosis, family history of psychosis, trauma index, executive function, processing speed, working memory, age, sex, race, ethnicity, and income-to-needs ratio. We selected these inputs in advance based on their availability in the ABCD dataset and their relevance as factors influencing schizophrenia or psychotic-like experiences. We entered variables using a simultaneous forced-entry strategy^41^, verified model stability using variance inflation factors to rule out collinearity, and evaluated model significance using Satterthwaite’s approximation.

For both models we transformed continuous inputs (i.e., trauma index, age, income-to-needs ratio, and measures of executive function, processing speed, or working memory) by dividing by two standard deviations, while leaving binary inputs untransformed. This scaling method has been recommended^42,43^ for situations where models include a mixture of continuous and binary inputs, because it allows regression coefficients to be directly compared when assessing relative effect sizes. This is because the difference between two standard-deviation changes for a continuous variable represents the change from a very low (∼2nd percentile) to a very high (∼98th percentile) value. Such a change is comparable to the difference between the two conditions (0 or 1) of a binary variable^42^.

As an additional exploratory analysis related to hypothesis 1, we looked for group differences in mean item-level PQ-BC distress scores between participants with and without ASD. We conducted multiple univariate analysis of variance (ANOVA) tests, again accounting for multiple testing via the Benjamini-Hochberg method.

To compare executive function, working memory, and processing speed among ASD children with psychotic-like experiences, ASD children without psychotic-like experiences, and children with psychotic-like experiences but not ASD (hypothesis 2), we derived three groups (hereafter referred to, for simplicity, as ASD+/PLS+, ASD+/PLS-, and ASD-/PLS+), again using the PQ-BC distress score cutoff of ≥ 2 standard deviations above the mean to represent the presence of likely significant psychotic-like experiences. We compared the three groups along each measure using one-way ANOVA, with Tukey’s post-hoc test to identify pairwise differences.

Last, to investigate the validity of our method of ASD ascertainment, we estimated a forced-entry logistic regression model with parent-reported ASD as the outcome. Sex, history of speech delay, maternal age, paternal age, race, ethnicity, age, and income-to-needs ratio were inputs. Given the limited number of ASD cases in our sample, we did not use a multilevel model here. Instead, we used generalized estimating equations to adjust for family unit^44^, and did not adjust for ABCD recruitment site.

For all analyses we set an *a priori* significance threshold of *α* = 0.05.

### 2.4 Software and data

We conducted analyses in R 3.5.1^45^ using lme4 1.1-21 for modeling.

Scripts to reproduce the results reported here are available from the authors at **https://github.com/amandeepjutla/2019-abcd-asd**.

The ABCD dataset is available to interested researchers through the NIMH Data Archive (**https://nda.nih.gov/**). As the ABCD dataset grows and changes over time, individual data releases are assigned permanent digital object identifiers (DOIs). We used release 2.0.1, which can be found at **http://dx.doi.org/10.15154/1504041**.

In deriving income-to-needs ratios, we used annual poverty threshold data for 2016, 2017, and 2018 from the US Census Bureau (**https://www.census.gov/**).

### 3 Results

#### 3.1 Sample characteristics

As shown in **Table 1**, the mean age of our sample’s 9,130 participants was 9.91 years (*SD* 0.62, range 9.00 – 10.92). 4,778 (52.33%) were male. Mean income-to-needs ratio was 4.51 (*SD* 2.88, range 0.10 – 16.02). Mean number of reported traumatic experiences (i.e., trauma index score) was 0.52 (*SD* 1.08, range 0 – 17). 957 participants (10.48%) had a parent-reported family history of psychosis.

**Table 1:** Characteristics of study participants across ASD and non-ASD groups.

Characteristics of study participants across ASD and non-ASD groups
| Variable | Ill sample (n = 9130) | No ASD (n = 8979) | ASD (n = 151) | p |
| --- | --- | --- | --- | --- |
| Continuous | *M (SD)* | *M (SD)* | *M (SD)* | *t-test* |
| Age (in years) | 9.91 (0.62) | 9.91 (0.62) | 10.00 (0.61) | 0.12 |
| **PQ-BC total score** | **2.56 (3.51)** | **2.53 (3.49)** | **4.00 (4.54)** | **1.18 × 10<sup>-3\*\*</sup>** |
| **PQ-BC distress score** | **3.50 (7.31)** | **3.46 (7.26)** | **5.99 (9.72)** | **7.24 × 10<sup>-3\*\*</sup>** |
| Income-to-needs ratio | 4.51 (2.88) | 4.51 (2.88) | 4.33 (3.03) | 0.48 |
| Trauma index score | 0.52 (1.08) | 0.52 (1.09) | 0.57 (0.78) | 0.45 |
| **NIH TB Card Sort (executive function)** | **97.38 (15.25)** | **97.43 (15.22)** | **94.29 (16.86)** | **0.04<sup>\*</sup>** |
| **NIH TB Pattern Comparison (processing sp)** | **94.18 (21.97)** | **94.25 (21.94)** | **89.66 (22.93)** | **0.04<sup>\*</sup>** |
| **NIH TB List Sorting (working memory)** | **101.57 (14.69)** | **101.64 (14.65)** | **97.44 (16.43)** | **0.01<sup>\*</sup>** |
| Maternal age in years at birth | 29.73 (6.05) | 29.72 (6.04) | 30.44 (6.34) | 0.20 |
| **Paternal age in years at birth** | **31.98 (6.90)** | **31.95 (6.89)** | **33.28 (7.08)** | **0.04<sup>\*</sup>** |
| Categorical | *Count (%)* | *Count (%)* | *Count (%)* | *χ<sup>2</sup>* |
| **Male sex** | **4778 (52.33)** | **4647 (51.75)** | **131 (86.75)** | **6.49 × 10<sup>-19\*\*\*</sup>** |
| White race | 6186 (67.76) | 6084 (67.76) | 102 (67.55) | 1 |
| Black race | 1185 (12.98) | 1166 (12.99) | 19 (12.58) | 1 |
| "Other" race | 1715 (18.78) | 1685 (18.77) | 30 (19.87) | 1 |
| Latinx ethnicity | 1708 (18.71) | 1686 (18.78) | 22 (14.57) | 0.48 |
| Family history of psychosis | 957 (10.48) | 937 (10.44) | 20 (13.25) | 0.49 |
| **History of speech delay** | **1535 (16.81)** | **1445 (16.09)** | **90 (59.60)** | **4.73 × 10<sup>-32\*\*\*</sup>** |

Note: *M* = mean; *SD* = standard deviation; *p*-values are Benjamini-Hochberg corrected.
\*: *p* < 0.05
\*\*: *p* < 0.01
\*\*\*: *p* < 0.001

Regarding race, 6,186 (67.75%) were white, 1,185 (12.98%) black, and 1,715 (18.78%) “other.” Of participants in the “other” category, 46 (0.50%) were Chinese, 30 (0.33%) Filipinx, 44 (0.48%) Indian, 6 (0.07%) Japanese, 9 (0.10%) Korean, 7 (0.08%) Vietnamese, 32 (0.35%) Native American, 1 (0.01%) Samoan, 18 (0.20%) “other Asian,” 8 (0.09%) “other Pacific islander,” 414 (4.53%) “other race,” 41 (0.45%) “unknown race,” and 1,141 (12.50%) multiracial. 1,708 (18.71%) participants were of Latinx ethnicity. Of Latinx participants, 971 (56.85%) were white, 68 (3.98%) were black, and 669 (39.17%) were “other.”

Mean PQ-BC total score was 2.56 (*SD* 3.51, range 0 – 20). Mean PQ-BC distress score was 3.50 (*SD* 7.31, range 0 – 84). 425 (4.65%) participants had a PQ-BC distress score ≥ the 2 standard deviations above the mean threshold (which meant a score above 18). 151 participants (1.65%) had a parent-reported ASD diagnosis.

Participants with ASD differed from those without it in several ways. They had higher PQ-BC total scores, *t*(153.00) = −3.95, *p* = 1.18 × 10^−3^ and higher distress scores, *t*(152.82) = −3.18, *p* = 7.24 × 10^−3^. Their executive function was relatively impaired, *t*(154.14) = 2.27, *p* = 0.04, as were processing speed, *t*(154.66) = 2.44, *p* = 0.04, and working memory, *t*(154.04) = 3.12, *p* = 0.01. They had older fathers, *t*(154.82) = −2.29, *p* = 0.04 and were more likely to be male, χ^2^ = 72.93, *p* = 4.70 × 10^−17^) or to have a history of speech delay, χ^2^= 201.01, *p* = 8.82 × 10^−45^.

#### 3.2 Hypothesis 1: ASD as predictor of psychotic-like experiences

##### 3.2.1 Linear model: ASD as predictor of continuous PQ-BC distress score

In the linear model, ASD had a strong positive effect on PQ-BC distress score (**Table 2**), with ASD youth having scores 2.46 points higher on average than non-ASD youth (*β* = 2.46, 95% confidence interval (CI) 1.32 – 3.60, *t* = 4.23, *p* = 2.31 × 10^−5^

**Table 2:** Predictors of continuous PQ-BC distress score.

| Predictor | $\beta$ | 95% CI | | SE | *t* | *p* |
| --- | --- | --- | --- | --- | --- | --- |
|  |  | Lower | Upper |  |  |  |
| **ASD** | **2.46** | **1.32** | **3.60** | **0.58** | **4.23** | **$2.31 \times 10^{-5***}$** |
| **Family history of psychosis** | **1.05** | **0.56** | **1.53** | **0.25** | **4.20** | **$2.76 \times 10^{-5***}$** |
| **Latinx ethnicity** | **0.99** | **0.52** | **1.45** | **0.24** | **4.19** | **$2.86 \times 10^{-5***}$** |
| **Black race** | **0.89** | **0.30** | **1.48** | **0.30** | **2.95** | **$3.17 \times 10^{-3**}$** |
| **Trauma index score** | **0.55** | **0.26** | **0.85** | **0.15** | **3.68** | **$2.30 \times 10^{-4***}$** |
| Male sex | 0.13 | -0.17 | 0.42 | 0.15 | 0.84 | 0.40 |
| NIH TB Pattern Comparison (processing speed) | -0.24 | -0.56 | 0.08 | 0.16 | -1.45 | 0.15 |
| NIH TB Card Sort (executive function) | -0.25 | -0.58 | 0.07 | 0.17 | -1.52 | 0.13 |
| White race | -0.37 | -0.78 | 0.03 | 0.21 | -1.80 | 0.07 |
| **Age** | **-0.75** | **-1.05** | **-0.46** | **0.15** | **-5.01** | **$5.43 \times 10^{-7***}$** |
| **NIH TB List Sorting (working memory)** | **-1.12** | **-1.43** | **-0.81** | **0.16** | **-7.11** | **$1.29 \times 10^{-12***}$** |
| **Income-to-needs ratio** | **-1.23** | **-1.58** | **-0.89** | **0.18** | **-7.02** | **$2.34 \times 10^{-12***}$** |
| (Intercept) | 3.25 | 2.55 | 3.95 | 0.36 | 9.07 | $1.68 \times 10^{-11***}$ |

Note: 95% CI = 95% Wald confidence interval; *p*-values estimated via Satterthwaite's approximation.\*: $p < 0.05$ \*\*: $p < 0.01$ \*\*\*: $p < 0.001$

This absolute effect size was at least as strong as those of other predictors in the model, including the positive predictor family history of psychosis, which increased PQ-BC distress scores by 1.05 points on average (*β* = 1.05, 95% CI 0.56 – 1.53, *t* = 4.20, *p* = 2.76 × 10^−5^), the positive predictor Latinx ethnicity, which increased scores by an average of 0.99 points, (*β* = 0.99, 95% CI 0.52 – 1.45, *t* = 4.19, *p* = 2.86 × 10^−5^the positive predictor black race, which increased scores by an average of 0.89 points (*β* = 0.89, 95% CI 0.30 – 1.48, *t* = 2.95, *p* = 3.17 × 10^−3^), the negative predictor income-to-needs ratio, for which an increase of two standard deviations decreased PQ-BC distress scores by an average of 1.23 points (*β* = −1.23, 95% CI -1.58 – -0.89, *t* = −7.02, *p* = 2.34 × 10^−12^), and the negative predictor working memory skills, for which a two-standard deviation increase decreased PQ-BC distress scores by an average of 1.12 points (*β* = −1.12, 95% CI -1.43 – -0.81, *t* = −7.11, *p* = 1.29 × 10^−12^).

The effect of ASD was also larger than the positive predictor trauma, for which an increase in trauma index score by two standard deviations raised PQ-BC scores by 0.55 points (*β* = 0.55, 95% CI 0.26 – 0.85, *t* = 3.68, *p* = 2.30 × 10^−4^) and the negative predictor age, for which an increse in two standard deviations reduced PQ-BC scores by an average of 0.75 points (*β* = −0.75, 95% C -1.05 – -0.46, *t* = −5.01, *p* = 5.43 × 10^−7^).

Sex, white race, executive functioning, and processing speed were not significant predictors in this model.

##### 3.2.2 Logistic model: ASD as predictor of PQ-BC score above a threshold

The logistic model, in we regressed against PQ-BC distress score ≥ 2 standard deviations above the mean (**Table 3**) illustrates ASD’s effect size in concrete terms. ASD predicted of PQ-BC score above this threshold (OR 3.18, 95% CI 1.77 – 5.69, *p* = 1.03 × 10^−4^) with an effect at least as strong as those of Latinx ethnicity (OR 1.69, 95% CI 1.26 – 2.26, *p* = 4.92 × 10^−4^), family history of psychosis (OR 1.68, 95% CI 1.25 – 2.25, *p* = 6.07 × 10^−4^), income-to-needs ratio (OR 0.49, 95% CI 0.38 – 0.64, *p* = 1.32 × 10^−7^), and working memory (OR 0.51, 95% CI 0.40 – 0.65, *p* = 2.70 × 10^−8^The effect of ASD was also stronger than those of trauma (OR 1.20, 95% CI 1.01 – 1.44, *p* = 4.25 × 10^−2^) and age (OR 0.75, 95% CI 0.60 – 0.94, *p* = 1.22 × 10^−2^). Sex, white race, black race, executive functioning, and processing speed were not significant predictors in this model.

**Table 3:** Predictors of PQ-BC distress score above 2 SD cutoff.

| Predictor | $\beta$ | 95% CI | | SE | z | p |
| --- | --- | --- | --- | --- | --- | --- |
|  |  | Lower | Upper |  |  |  |
| **ASD** | **3.18** | **1.77** | **5.69** | **0.94** | **3.88** | **$1.03 \times 10^{-4***}$** |
| **Latinx ethnicity** | **1.69** | **1.26** | **2.26** | **0.25** | **3.48** | **$4.92 \times 10^{-4***}$** |
| **Family history of psychosis** | **1.68** | **1.25** | **2.25** | **0.25** | **3.43** | **$6.07 \times 10^{-4***}$** |
| Black race | 1.24 | 0.87 | 1.76 | 0.22 | 1.17 | 0.24 |
| **Trauma index score** | **1.20** | **1.01** | **1.44** | **0.11** | **2.03** | **0.04\*** |
| NIH TB Card Sort (executive function) | 0.96 | 0.75 | 1.23 | 0.12 | -0.31 | 0.76 |
| Male sex | 0.96 | 0.78 | 1.19 | 0.10 | -0.38 | 0.71 |
| NIH TB Pattern Comparison (processing spee | 0.88 | 0.70 | 1.11 | 0.10 | -1.10 | 0.27 |
| White race | 0.79 | 0.60 | 1.04 | 0.11 | -1.67 | 0.09 |
| **Age** | **0.75** | **0.60** | **0.94** | **0.08** | **-2.51** | **0.01\*** |
| **NIH TB List Sorting (working memory)** | **0.51** | **0.40** | **0.65** | **0.06** | **-5.56** | **$2.70 \times 10^{-8***}$** |
| **Income-to-needs ratio** | **0.49** | **0.38** | **0.64** | **0.07** | **-5.28** | **$1.32 \times 10^{-7***}$** |
| (Intercept) | 0.02 | 0.01 | 0.04 | 0.01 | -14.34 | $1.23 \times 10^{-46***}$ |

Note: 95% CI = 95% Wald confidence interval; *p*-values estimated via Satterthwaite's approximation.\*: $p < 0.05$ \*\*: $p < 0.01$ \*\*\*: $p < 0.001$

##### 3.2.3 Exploratory analysis: group differences in item-level PQ-BC distress scores

The results of univariate ANOVAs comparing mean item-level PQ-BC distress scores between participants with and without ASD are provided in **Table S2**. After Benjamini-Hochberg correction, individuals with ASD scored higher than those without on eleven items.

In ascending order of statistical significance, these were items #14 (“Did you feel confused because something you experienced didn’t seem real, or it seemed imaginary to you?”, F(1, 9,128 = 5.20, *p* = 0.04)), #12 (“Did you start to worry at times that your mind was trying to trick you or was not working right?”, F(1, 9,128 = 6.27, *p* = 0.03)), #20 (“Did you suddenly start to be able to see things that other people could not see, or they did not seem to see?”, F(1, 9,128 = 6.04, *p* = 0.03)), #1 (“Did places that you know well, such as your bedroom, or other rooms in your home, your classroom, or school yard, suddenly seem weird, strange or confusing to you, like not the real world?”, F(1, 9,128 = 6.38, *p* = 0.03)), #2 (“Did you hear strange sounds that you never noticed before, like banging, clicking, hissing, clapping, or ringing in your ears?”, F(1, 9,128 = 7.30, *p* = 0.02)), #10 (“Did you lose concentration because you noticed sounds in the distance that you usually don’t hear?”, F(1, 9,128 = 8.00, *p* = 0.02)), #17 (“Did you feel that sometimes your thoughts were strong you could almost hear them, as if another person, not you, spoke them?”, F(1, 9,128 = 7.21, *p* = 0.02)), #18 (“Did you feel that other people might want something bad to happen to you or that you could not trust other people?”, F(1, 9,128 = 7.08, *p* = 0.02)), #11 (“Although you could not see anything or anyone, did you suddenly start to feel that an invisible energy, creature, or some person was around you?”, F(1, 9,128 = 10.80, *p* = 0.01)), #15 (“Did you honestly believe in things that other people would say are unusual or weird?”, F(1, 9,128 = 11.29, *p* = 0.01)), and #21 (“Did you suddenly start to notice that people sometimes had a hard time understanding what you were saying, even though they used to understand you well?”, F(1, 9,128 = 13.65, *p* = 4.65 × 10^−3^)).

### 3.3 Hypothesis 2: Different neurocognitive profiles among sub-groups

Our results did not support hypothesis 2, that neurocognitive profiles would differ between ASD+/PLS+ (n = 18), ASD+/PLS- (n = 133) and ASD-/PLS+ (n = 407) participants. We did not identify significant differences between these groups along any of the three NIH Toolbox measures we had selected (executive function: F(2, 555 = 1.28, *p* = 0.28); processing speed: F(2, 555 = 0.15, *p* = 0.86); working memory: F(2, 555 = 2.27, *p* = 0.10).

#### 3.4 Robustness of findings

We tested the robustness of our findings by addressing a notable limitation of the approach we took: namely, that our ascertainment of ASD was based on parent report, and not on gold-standard diagnosis by a clinician. This necessitated that we assume parent-reported ASD was a reasonably accurate proxy for “confirmed” ASD.

To examine this assumption, we estimated a logistic regression model with parent-reported ASD as the outcome and a series of predictors chosen based on their having a previously-described association with ASD and their availability in the ABCD dataset: male sex, history of speech delay, paternal age, and maternal age^1^. We also included race, ethnicity, age, and income-to-needs as covariates. The results of this regression, shown in **Table S3**, indicate that the predictors of parent-reported ASD in the ABCD cohort are consistent with known predictors of ASD in the general population, with history of speech delay (OR 6.09, 95% CI 4.34 – 8.53, *p* < 2 × 10^−16^), male sex (OR 4.48, 95% CI 2.77 – 7.25, *p* = 1.06 × 10^−9^) and paternal age (OR 1.64, 95% CI 1.02 – 2.64, *p* = 0.04) reaching statistical significance. We consider these results an indication that our use of parent-reported ASD in this study is reasonable.

#### 3.4 Sensitivity analyses

In sensitivity analyses, we estimated alternate versions of the models used to test hypothesis 1. We found that the pattern of our results was robust when we restricted our analysis to households with only one child participating in ABCD and when we used PQ-BC total score rather than distress score as our dependent variable. In both cases, ASD’s effect remained at least as strong as those of other predictors in the model.

## 4 Discussion

Our finding that ASD diagnosis predicts psychotic-like experiences in youth is consistent with literature suggesting that rates of SCZ and psychosis are greater in adults with ASD than in the general population^14,15^. It also adds new information in suggesting that the magnitude of ASD as a risk factor for psychotic-like experiences is significant even when compared to other established SCZ risk factors, such as family history and lower income-to-needs ratio.

Although it is possible that ASD is acting as a proxy for another SCZ risk factor we have not considered, we consider this unlikely, as it is unclear what this factor would be. In contrast, black race and Latinx ethnicity, which in our results also appeared to predict psychotic-like experiences, could have been proxies for other known SCZ/psychosis risk factors not included in our models, such as urbanicity, an ethnic density effect, or first- or second-generation immigration status^46^.

Our results therefore may have implications for both clinicians working primarily with ASD youth and clinicians working with CHR and early psychosis populations. Clinicians who see adolescents with ASD may benefit from incorporating psychosis screens into their assessments, and referring to CHR assessment clinics when indicated. By the same token, clinicians working with the CHR population might consider finding ways to connect with those working with the ASD population to ensure that a referral pipeline is in place.

Findings also are of import for researchers interested in the nosology of ASD and psychotic disorders. Our exploratory finding that ASD youth had higher mean distress scores on 11 of the PQ-BC’s 21 items requires independent replication, but has interesting implications. Although some of these items may indicate individual psychotic-like pheomena of particular importance in the ASD population, some could simply reflect the sensory hypersensitivity, special or unusual interests, and communication difficulties that are common in ASD^1^. The possibility that some questions are indexing core ASD features rather than unique psychotic-like symptoms highlights the difficulty of identifying psychotic experiences in ASD. It suggests a possible need for adapted assessment instruments that capture the unique phenomenology of psychosis in this population.

We were not able to identify a cognitive profile specific to individuals having both reported ASD and psychotic-like experiences. This may in part be a function of sample size. Although the ABCD cohort is large, only 151 participants had reported ASD, and of those, 18 also had psychotic-like symptoms. Future studies that combine data collected from multiple large cohorts might have the power to detect subtle differences along cognitive dimensions. In the future, studies might also explore aspects of social cognition, language, and sensory functioning to better dissociate these groups.

We acknowledge that our study has some important limitations. Our method of ASD ascertainment – parent report of an ASD diagnosis – is less robust than a gold-standard diagnosis might have been. However, evidence suggests that parent-reported ASD is often accurate. In one study of 118 youth with parent-reported ASD enrolled in the nationwide Interactive Autism Network registry, parents were able to provide records confirming an ASD diagnosis in 116 (98.31%) cases^47^. In a similar study of enrollees in the Autism Spectrum Database – UK study, records substantiated parent-reported ASD in 142 of 156 (91.03%) cases^48^. Our finding that predictors of parent-reported ASD in the ABCD cohort are similar to known predictors of ASD lends further support to the idea that parent report is a meaningful indicator.

Although ABCD is not an ASD-focused study, it is notable that even in this dataset, collected without an intention to examine ASD as a risk factor for psychotic-like experiences, we nevertheless found a strong relationship. In our view, this speaks to the robustness of this association. It also supports results from registry studies that similarly were not designed with an eye towards recruiting participants with ASD or psychotic symptoms^49,50^.

The ABCD cohort is relatively young, and most work examining convergence between ASD and schizophrenia has been conducted with older groups who have formal diagnoses. This limits the generalizability of our findings. However, our identification of an association between ASD and psychotic-like symptoms well before the psychosis prodrome typically appears raises intriguing possibilities for future research.

This is, to our knowledge, the first study to examine the association between ASD and psychotic-like experiences in middle childhood. As the ABCD cohort ages, and as longitudinal data is collected, we hope to delineate how ASD relates to trajectories of psychotic-like experiences over time, leading to a more robust understanding of how, when, and why ASD and SCZ converge.

## Data Availability

The Adolescent Brain Cognitive Development (ABCD) cohort dataset is available to interested researchers through the NIMH Data Archive.

https://nda.nih.gov/

http://dx.doi.org/10.15154/1504041

https://github.com/amandeepjutla/2019-abcd-asd

## Data Availability

https://nda.nih.gov/

http://dx.doi.org/10.15154/1504041

https://github.com/amandeepjutla/2019-abcd-asd

## 5 Acknowledgements

This project was financially supported by a Whitaker Scholar in Developmental Neuropsychiatry Award to Dr. Jutla funded by the Marilyn and James Simons Foundation.

The ABCD study is supported by the National Institutes of Health and additional federal partners under award numbers U01DA041022, U01DA041028, U01DA041048, U01DA041089, U01DA041106, U01DA041117, U01DA041120, U01DA041134, U01DA041148, U01DA041156, U01DA041174, U24DA041123, and U24DA041147. Information about the study’s supporters, participating sites, and study investigators can be found at https://abcdstudy.org. Although ABCD investigators provided data, they did not participate in the analysis or writing of this report. This manuscript therefore reflects the views of the authors and does not necessarily reflect the opinions or views of the NIH or ABCD consortium investigators.

## Supplementary Tables

**Table S1:**
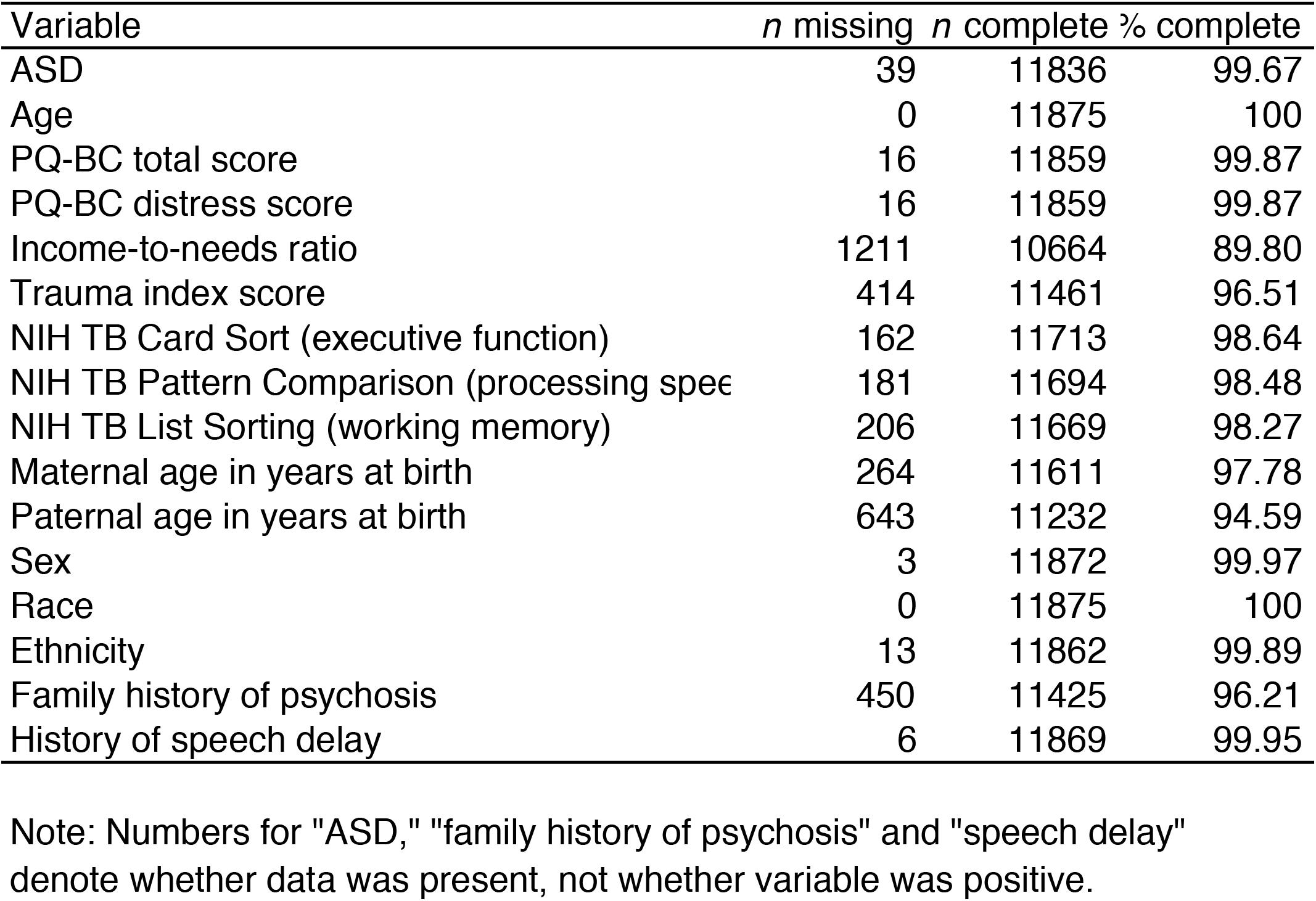
Missingness by variable.

**Table S2:** Item-level PQ-BC distress score comparisons.

| PQ-BC item |  | *M (SD)* |  | *F* | *p* |
| --- | --- | --- | --- | --- | --- |
| Number | Question ("In the past month...") | No ASD (n = 8979) | ASD (n = 151) |  |  |
| 1 | **Did places that you know well, such as your bedroom, or other rooms in your home, your classroom, or school yard, suddenly seem weird, strange or confusing to you, like not the real world?** | **0.08 (0.50)** | **0.19 (0.77)** | **6.38** | **0.03\*** |
| 2 | **Did you hear strange sounds that you never noticed before, like banging, clicking, hissing, clapping, or ringing in your ears?** | **0.30 (0.88)** | **0.50 (1.20)** | **7.30** | **0.02\*** |
| 3 | Did things you looked at seem different than they usually do, like did they seem shinier or darker, larger or smaller, or changed in some other way? | 0.08 (0.47) | 0.09 (0.52) | 0.03 | 0.86 |
| 4 | Did you feel like you had special, unusual powers, like you could make things happen by magic, or that you could magically know what was inside another person's mind, or magically know what was going to happen in the future when other people could not? | 0.04 (0.36) | 0.05 (0.27) | 0.03 | 0.86 |
| 5 | Did you feel that someone else, who is not you, has taken control over the private, personal thoughts or ideas inside your head? | 0.11 (0.61) | 0.21 (0.87) | 4.23 | 0.07 |
| 6 | Did you suddenly find it hard to figure out how to say something quickly and easily so that other people would understand what you meant? | 0.16 (0.64) | 0.21 (0.74) | 1.01 | 0.37 |
| 7 | Did you ever feel very certain that you have very special abilities or magical talents that other people do not have? | 0.03 (0.30) | 0.07 (0.50) | 2.40 | 0.16 |
| 8 | Did you suddenly feel that you could not trust other people because they seemed to be watching you or talking about you in an unfriendly way? | 0.45 (1.11) | 0.60 (1.37) | 2.81 | 0.13 |
| 9 | Did your skin or just beneath your skin suddenly start feeling strange, like bugs crawling? | 0.26 (0.88) | 0.34 (1.03) | 1.13 | 0.36 |
| 10 | **Did you lose concentration because you noticed sounds in the distance that you usually don't hear?** | **0.22 (0.81)** | **0.41 (1.14)** | **8.00** | **0.02\*** |
| 11 | **Although you could not see anything or anyone, did you suddenly start to feel that an invisible energy, creature, or some person was around you?** | **0.38 (1.09)** | **0.68 (1.49)** | **10.80** | **0.01\*** |
| 12 | **Did you start to worry at times that your mind was trying to trick you or was not working right?** | **0.20 (0.81)** | **0.36 (1.12)** | **6.27** | **0.03\*** |
| 13 | Did you feel that the world is not real, you are not real, or that you are dead? | 0.09 (0.57) | 0.18 (0.83) | 3.66 | 0.09 |
| 14 | **Did you feel confused because something you experienced didn't seem real, or it seemed imaginary to you?** | **0.11 (0.58)** | **0.22 (0.84)** | **5.20** | **0.04\*** |
| 15 | **Did you honestly believe in things that other people would say are unusual or weird?** | **0.12 (0.63)** | **0.29 (1.01)** | **11.29** | **0.01\*** |
| 16 | Did you feel that parts of your body had suddenly changed or worked differently than before, like your legs had suddenly turned to something else, or your nose could suddenly smell things you'd never actually smelled before? | 0.06 (0.43) | 0.07 (0.34) | 0.09 | 0.84 |
| 17 | **Did you feel that sometimes your thoughts were so strong you could almost hear them, as if another person, NOT you, spoke them?** | **0.09 (0.53)** | **0.21 (0.81)** | **7.21** | **0.02<sup>\*</sup>** |
| 18 | **Did you feel that other people might want something bad to happen to you or that you could not trust other people?** | **0.28 (0.95)** | **0.49 (1.28)** | **7.08** | **0.02<sup>\*</sup>** |
| 19 | Did you suddenly start to see unusual things that you never saw before, like flashes, flames, blinding light, or shapes floating in front of you? | 0.17 (0.76) | 0.28 (0.95) | 3.06 | 0.12 |
| 20 | **Did you suddenly start to be able to see things that other people could not see, or they did not seem to see?** | **0.11 (0.63)** | **0.24 (0.88)** | **6.04** | **0.03<sup>\*</sup>** |
| 21 | **Did you suddenly start to notice that people sometimes had a hard time understanding what you were saying, even though they used to understand you well?** | **0.13 (0.62)** | **0.32 (1.06)** | **13.65** | **4.65 × 10<sup>-3\*\*</sup>** |

Note: *M* = mean; *SD* = standard deviation; *p*-values are Benjamini-Hochberg corrected.
<sup>\*</sup>: *p* < 0.05
<sup>\*\*</sup>: *p* < 0.01

**Table S3:**
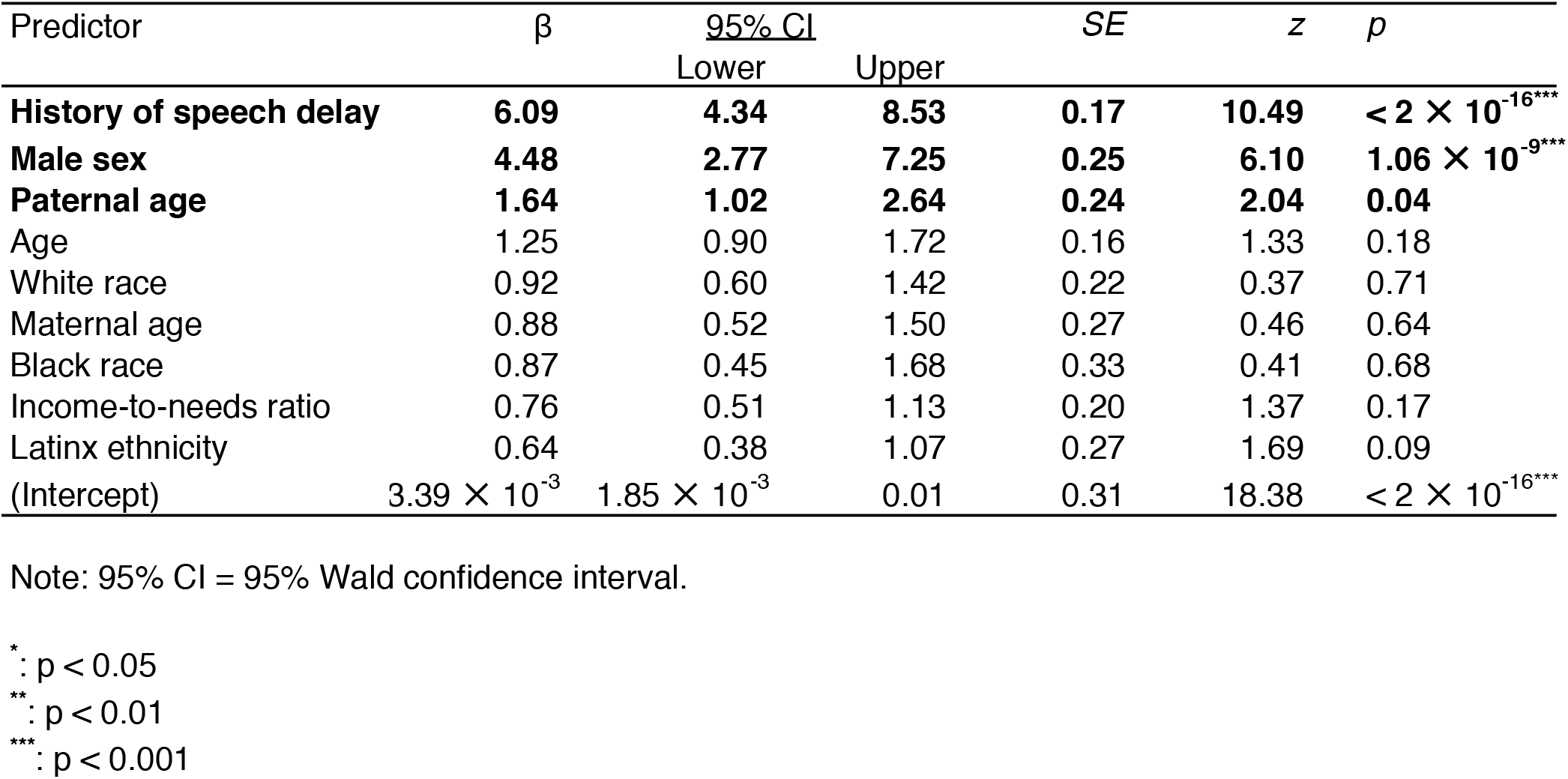
Predictors of parent-reported ASD.

| Predictor | $\beta$ | 95% CI | | SE | z | p |
| --- | --- | --- | --- | --- | --- | --- |
|  |  | Lower | Upper |  |  |  |
| **History of speech delay** | **6.09** | **4.34** | **8.53** | **0.17** | **10.49** | **$< 2 \times 10^{-16}$**<sup>***</sup> |
| **Male sex** | **4.48** | **2.77** | **7.25** | **0.25** | **6.10** | **$1.06 \times 10^{-9}$**<sup>***</sup> |
| **Paternal age** | **1.64** | **1.02** | **2.64** | **0.24** | **2.04** | **0.04** |
| Age | 1.25 | 0.90 | 1.72 | 0.16 | 1.33 | 0.18 |
| White race | 0.92 | 0.60 | 1.42 | 0.22 | 0.37 | 0.71 |
| Maternal age | 0.88 | 0.52 | 1.50 | 0.27 | 0.46 | 0.64 |
| Black race | 0.87 | 0.45 | 1.68 | 0.33 | 0.41 | 0.68 |
| Income-to-needs ratio | 0.76 | 0.51 | 1.13 | 0.20 | 1.37 | 0.17 |
| Latinx ethnicity | 0.64 | 0.38 | 1.07 | 0.27 | 1.69 | 0.09 |
| (Intercept) | $3.39 \times 10^{-3}$ | $1.85 \times 10^{-3}$ | 0.01 | 0.31 | 18.38 | **$< 2 \times 10^{-16}$**<sup>***</sup> |

Note: 95% CI = 95% Wald confidence interval.
\*: $p < 0.05$
\*\*: $p < 0.01$
\*\*\*: $p < 0.001$

